# The impact of neighborhood socioeconomic deprivation on metastatic pancreatic cancer treatment and survival: An incidence-based, causally-structured observational study

**DOI:** 10.64898/2026.08.06.26359821

**Authors:** Apurva Raghu, Shreya Shah, Aishwarya Pattnaik, Jennifer B. Permuth, Margaret Park, Hejer Dhahri, Huang Chiao Huang, Jason B. Fleming, Daniel A. Anaya, Benjamin D. Powers

## Abstract

**Purpose:** Metastatic pancreatic ductal adenocarcinoma (PDAC) portends a poor prognosis. Prior studies have assessed the association of socioeconomic deprivation (SED) in PDAC often with large geographic areas. This study employed a causal framework to characterize neighborhood SED on treatment receipt and survival in metastatic PDAC.

**Methods:** Using the incidence-based Florida Cancer Data System, metastatic PDAC patients diagnosed from 2007-2015 were identified. The Area Deprivation Index, a composite measure of SED that ranks neighborhoods from 1-100 (higher scores = higher deprivation), was used to assess receipt of systemic therapy and overall survival (OS). Exposures and covariates were assessed using descriptive statistics and a causal inference framework.

**Results:** Overall, 9,574 patients met inclusion criteria. 46.6% of patients received systemic therapy, ranging 39.4% to 54% in the highest and lowest SED quartiles, respectively. After adjustment, the lowest quartile had increased odds of systemic therapy relative to the highest (OR 1.93; 95% CI 1.70-2.18). Median OS was 3.8 months for the lowest quartile and 2.4 months for the highest (p = 0.01). Patients in the highest quartile had an estimated 32% higher hazard of death than the lowest (HR 1.32, 95% bootstrap CI 1.20-1.40).

**Conclusion:** In an incidence-based statewide cohort, most patients did not receive treatment for metastatic PDAC and median OS was poor—2.9 months. Using a causal inference framework, higher SED led to lower rates of systemic therapy receipt and worse overall survival in metastatic PDAC. Future research should focus on the mechanisms that shape these findings.

**Context Summary:** *Key objective:* Determine the effect of neighborhood socioeconomic deprivation on treatment receipt and survival in metastatic pancreatic ductal adenocarcinoma (PDAC) using a causally structured analysis.

*Knowledge generated:* In this retrospective cohort study of 9,574 patients, 46.6% received treatment and median OS was poor—2.9 months. Using a causal inference framework, the highest socioeconomic deprivation quartile received less systemic treatment and had a 32% increased hazard of death relative to the lowest quartile.

*Relevance:* This study provides evidence of a causal effect of socioeconomic deprivation on systemic treatment and overall survival in metastatic PDAC. These findings provide a basis for future studies to define and sculpt interventions to remedy the impact of neighborhood socioeconomic disparities on metastatic PDAC.

## Introduction

Pancreatic ductal adenocarcinoma (PDAC) is projected to become the second leading cause of cancer-related death by 2030.^1^ Roughly 50-55% of patients present with metastatic disease.^2^ The standard of care for metastatic PDAC is systemic therapy.^3^ Despite advances in chemotherapy regimens and promising results of clinical trials, population-based outcomes remain poor. The 5-year survival is 3% for metastatic disease, one of the lowest of all cancers, and the median overall survival is around 2 months.^4–6^ Even with improvements in multi-agent regimens, stage IV survival has only increased from 3.5 to 3.9 months from the 2000s to the 2010s, underscoring the need to better understand factors that influence receipt of treatment and survival.^7^

Social determinants of health have been reported to be associated with PDAC outcomes.^8,9^ Area-level socioeconomic deprivation has been linked to lower likelihood of receiving stage-appropriate therapy and higher mortality, potentially reflecting structural barriers such as geographic distance, and delayed or non-referral to high-volume centers.^10,11^ Disparities in receipt of therapy have also been observed in non-metastatic disease, including differences in utilization of adjuvant treatment.^12^

However, few studies have assessed the association of neighborhood-level deprivation on treatment receipt and survival in a metastatic PDAC cohort^13–15^ Additionally, given the poor outcome and short follow-up, it is unclear if socioeconomic status exerts an influence on treatment or survival in metastatic PDAC. To address these gaps in the literature, we conducted a retrospective cohort study using the Florida Cancer Database to causally estimate the effect of neighborhood socioeconomic deprivation on systemic therapy receipt and overall survival among patients with metastatic PDAC.

## Methods

### Study Setting and Cohort

A retrospective cohort study was conducted using incidence-based data from the Florida Cancer Data System (FCDS). Patients over the age of 18 diagnosed with stage IV PDAC between 2007 and 2015 were identified using the International Classification of Diseases for Oncology (ICD-O) and histology codes (8500/3 ductal adenocarcinoma NOS, 8140/3 adenocarcinoma NOS). Patients were excluded for non-adenocarcinoma, diagnosis by autopsy or death certificate only, no pathologic diagnosis, non-metastatic disease, or unknown Area Deprivation Index (ADI) (Supplemental Figure 1). This study was approved by the by the Advarra Institutional Review Board (MCC19853) and the Florida Department of Health IRB.

### Exposure Variable

The exposure variable was neighborhood socioeconomic deprivation, measured using the area deprivation index (ADI). The ADI is a validated, composite measure of socioeconomic deprivation that includes 17 different education, employment, housing quality, and poverty measures that have been weighted and summed to produce scores for each neighborhood (census block group) on a scale from 1 to 100 (lowest to highest deprivation) at the national level.^16^ It was created from US Census data and American Community Survey data and made publicly available via the Neighborhood Atlas.^17^ In line with prior studies, ADI was categorized into quartiles with similar total count size for ease of data interpretation.^18^

### Outcome Variables and Covariates

The outcomes were receipt of systemic therapy (including chemotherapy, immunotherapy, and targeted therapy), and overall survival. Receipt of treatment was analyzed as a binary variable with patients categorized as received or not received. Patients with unknown treatment status were excluded from regression analysis. Overall survival was defined as time from date of diagnosis to death date from any cause. Patients were censored at last known follow-up. Covariates included time to initiation of systemic treatment, which was defined as the time from date of diagnosis until the date of the first course of systemic treatment, age at diagnosis, gender, race, ethnicity, primary insurance payer at diagnosis, and tumor grade.

### Statistical Analysis

Continuous variables were reported as medians with interquartile ranges and categorical variables were reported as counts and proportions. Associations with socioeconomic deprivation quartiles were assessed using the Kruskal Wallis test for continuous variables and chi-square test for categorical variables. Overall survival distributions were estimated using the Kaplan-Meier method for the overall cohort and after stratification by receipt of systemic therapy.

The study objective was to identify the effect of neighborhood socioeconomic deprivation on receipt of systemic therapy and overall survival; therefore, a causal approach was used.^19,20^ To visualize causal relationships and provide a structure to identify confounders, mediators, and colliders, directed acyclic graphs (DAG) were constructed.^21–23^ This approach helps avoid biases, including confounding, overadjustment, and collider stratification bias and makes explicit the assumed relationships between variables.^21^ For this study, DAGitty.net website was used for minimal sufficient adjustment of covariates after visualization of exposures, outcomes, and other related variables to achieve causal estimates (Supplemental Figure 2).^24^

To estimate the effect of socioeconomic deprivation by quartile on receipt of systemic therapy (yes/no), logistic regression models were constructed with minimally sufficient adjustment for age, sex, race, and ethnicity and presented as odds ratios with 95% confidence intervals. A marginal plot was created based on the adjusted logistic regression model using socioeconomic deprivation as a continuous variable and the slope of the curve representing the marginal effect, defined as the change in probability of systemic therapy for each change in ADI value and are presented with 95% confidence intervals.^25^

To produce a causal estimate of the marginal total effect of neighborhood socioeconomic deprivation on overall survival, inverse probability weighting (IPW) with a generalized propensity score for the multinomial exposure was used.^26^ A multinomial logistic regression model was fit with socioeconomic deprivation quartiles as the outcome and age, race, ethnicity, and sex to produce the generalized propensity score, i.e., the conditional probability of belonging to their observed deprivation quartile given their measured confounders. Stabilized IPWs were then constructed as the ratio of the marginal probability of each patient’s observed quartile to their conditional probability from the propensity score model. Extreme weights were trimmed (at the 1st and 99th percentiles). The effective sample size after weighting was calculated for each quartile as the square of the sum of weights divided by the sum of squared weights. Covariate balance was assessed between each deprivation quartile and the reference quartile before and after weighting and quantified using the absolute standardized mean difference (ASMD) for each confounder. An ASMD below 0.10 after weighting was considered indicative of adequate balance and achieved for 12 of 13 confounders, except for Black race, for which the adjusted ASMD was 0.1005—marginally exceeding the pre-specified threshold. As the 0.10 threshold is widely adopted, it represents a convention rather than a statistically derived cutoff and simulations have suggested that ASMD exceedances of this magnitude produce negligible bias in IPW-estimated treatment effects.^27–29^

Marginal survival curves in the IPW-weighted pseudo-population were estimated using the Kaplan-Meier method with stabilized trimmed weights incorporated as frequency weights. Differences in weighted survival distributions across socioeconomic deprivation quartiles were assessed using weighted log-rank tests. To estimate the marginal hazard ratio for the effect of socioeconomic deprivation on overall survival, a weighted Cox proportional hazards model was fit with deprivation quartile as the exposure, incorporating stabilized trimmed weights as probability weights with robust variance estimation to account for the induced weighting. The proportional hazards assumption was evaluated using scaled Schoenfeld residual plots. All survival estimates and hazard ratios were derived using nonparametric bootstrap resampling (500 samples and 42 seeds).

To characterize the potential impact of unmeasured confounding, E-values were computed for the primary IPW-estimated hazard ratios. The E-value quantifies the minimum strength of association, on the risk ratio scale, that an unmeasured confounder would need to have with both the exposure and the outcome to fully explain the observed association.^30^ E-values were computed for both the point estimate and the lower bound of the 95% confidence interval.

Finally, a formal causal mediation analysis could not be performed as this would require additional assumptions regarding unmeasured confounding from comorbidity data that are not present in the dataset. Therefore, a non-causal, descriptive assessment of the potential mediation pathways through which socioeconomic deprivation may influence overall survival was performed.^31^ For this, three sequentially adjusted Cox models were estimated in the unweighted observed sample. Model 1 included socioeconomic deprivation quartiles and measured confounders only (i.e., the total effect estimates); Model 2 additionally adjusted for insurance status; and Model 3 with the addition of receipt of systemic therapy. All analyses were conducted using Stata statistical software (StataNow/SE 19.5; StataCorp LLC, College Station, TX) and R (version 4.6.0; R Foundation for Statistical Computing, Vienna, Austria) using the analytic packages: survival v3.8.6, ggsurvfit v1.2.0, ggplot2 v4.0.3, cobalt v4.6.3.^32^ Statistical tests were two-sided and p-value <0.05 was considered statistically significant.

## Results

### Study Cohort and Patient Characteristics

A total of 24,277 PDAC cases were identified and 9,574 met inclusion criteria. Overall, 53.8% patients were male and the median age at diagnosis was 70 years, (interquartile range 61-79). Across quartiles, there were statistically significant differences in age at diagnosis, race, ethnicity, and insurance (Table 1). African Americans constituted a higher proportion of cases in the highest (18.2%) compared to the lowest deprivation quartile (4.5%), respectively. Conversely, Hispanic/Latino ethnicity had a higher proportion of cases in lowest deprivation quartile (13.9%) compared to the highest (8.8%), respectively. Although private insurance was more represented in the lower deprivation quartile (25.8%) relative to the highest (18.9%), Medicare showed a small gradient from the lowest to highest quartiles (59-62%).

**Table 1:** Characteristics of Metastatic Pancreatic Adenocarcinoma by Socioeconomic Deprivation Quartile (ADI percentile score range)

|  | Q1 Lowest deprivation (1-35) | Q2 Low (36-57) | Q3 High (58-79) | Q4 Highest deprivation (80-100) | p-value |
| --- | --- | --- | --- | --- | --- |
| **Total** | 2,420 | 2,436 | 2,400 | 2,318 |  |
| **Age at Diagnosis** | 71.0 (63.0-79.0) | 70.0 (62.0-77.0) | 69.0 (61.0-78.0) | 70.0 (61.0-79.0) | <0.001 |
| **Gender** |  |  |  |  | 0.02 |
| Male | 1,345 (55.6) | 1,339 (55.0) | 1,302 (54.2) | 1,167 (50.3) |  |
| Female | 1,074 (44.4) | 1,096 (45.0) | 1,098 (45.8) | 1,149 (49.6) |  |
| Transexual | 0 (0.0%) | 0 (0.0%) | 0 (0.0%) | 1 (0.0%) |  |
| Unknown | 1 (0.0%) | 1 (0.0%) | 0 (0.0%) | 1 (0.0%) |  |
| **Race** |  |  |  |  | <0.001 |
| White | 2,262 (93.5%) | 2,189 (89.9%) | 1,995 (83.1%) | 1,860 (80.2%) |  |
| Black/African American | 110 (4.5%) | 174 (7.1%) | 348 (14.5%) | 422 (18.2%) |  |
| Asian/Pacific Islander | 24 (1.0%) | 33 (1.4%) | 22 (0.9%) | 13 (0.6%) |  |
| Other | 12 (0.5%) | 20 (0.8%) | 15 (0.6%) | 11 (0.5%) |  |
| Unknown | 12 (0.5%) | 20 (0.8%) | 20 (0.8%) | 12 (0.5%) |  |
| **Ethnicity** |  |  |  |  | <0.001 |
| Spanish-Hispanic-Latino | 336 (13.9%) | 329 (13.5%) | 316 (13.2%) | 204 (8.8%) |  |
| Non-Spanish-Hispanic-Latino | 2,072 (85.6%) | 2,096 (86.0%) | 2,068 (86.2%) | 2,104 (90.8%) |  |
| Unknown | 12 (0.5%) | 11 (0.5%) | 16 (0.7%) | 10 (0.4%) |  |
| **Primary Payer at Diagnosis** |  |  |  |  | <0.001 |
| Private insurance | 625 (25.8%) | 550 (22.6%) | 514 (21.4%) | 437 (18.9%) |  |
| Medicare | 1,475 (61.0%) | 1,457 (59.8%) | 1,409 (58.7%) | 1,429 (61.6%) |  |
| Medicaid | 69 (2.9%) | 138 (5.7%) | 178 (7.4%) | 194 (8.4%) |  |
| Military/TRICARE/Veterans Affairs | 38 (1.6%) | 44 (1.8%) | 68 (2.8%) | 40 (1.7%) |  |
| Insured, NOS | 77 (3.2%) | 90 (3.7%) | 78 (3.2%) | 74 (3.2%) |  |
| Uninsured | 75 (3.1%) | 99 (4.1%) | 112 (4.7%) | 79 (3.4%) |  |
| Unknown | 61 (2.5%) | 58 (2.4%) | 41 (1.7%) | 65 (2.8%) |  |
| **Grade** |  |  |  |  | 0.36 |
| 1 | 45 (1.9%) | 43 (1.8%) | 39 (1.6%) | 50 (2.2%) |  |
| 2 | 181 (7.5%) | 208 (8.5%) | 209 (8.7%) | 182 (7.9%) |  |
| 3 | 345 (14.3%) | 330 (13.5%) | 324 (13.5%) | 275 (11.9%) |  |
| 4 | 16 (0.7%) | 21 (0.9%) | 19 (0.8%) | 13 (0.6%) |  |
| Unknown | 1,833 (75.7%) | 1,834 (75.3%) | 1,809 (75.4%) | 1,798 (77.6%) |  |
| **Systemic Therapy** |  |  |  |  | <0.001 |
| None | 1,012 (41.8%) | 1,114 (45.7%) | 1,241 (51.7%) | 1,293 (55.8%) |  |
| Administered | 1,307 (54.0%) | 1,207 (49.5%) | 1,039 (43.3%) | 914 (39.4%) |  |
| Unknown | 101 (4.2%) | 115 (4.7%) | 120 (5.0%) | 111 (4.8%) |  |
| **Time to Initiation of Systemic Treatment (Months)** | 0.8 (0.5-1.3) | 0.8 (0.5-1.3) | 0.9 (0.5-1.4) | 0.9 (0.5-1.4) | 0.17 |
| **30-Day Mortality** |  |  |  |  | <0.001 |
| No | 1,959 (81.0%) | 1,929 (79.2%) | 1,836 (76.5%) | 1,722 (74.3%) |  |
| Yes | 461 (19.0%) | 507 (20.8%) | 564 (23.5%) | 596 (25.7%) |  |
| **90-Day Mortality** |  |  |  |  | <0.001 |
| No | 1,370 (56.6%) | 1,244 (51.1%) | 1,149 (47.9%) | 1,015 (43.8%) |  |
| Yes | 1,050 (43.4%) | 1,192 (48.9%) | 1,251 (52.1%) | 1,303 (56.2%) |  |
| **1-year Mortality** |  |  |  |  | <0.001 |
| No | 439 (18.1%) | 353 (14.5%) | 295 (12.3%) | 238 (10.3%) |  |
| Yes | 1,981 (81.9%) | 2,083 (85.5%) | 2,105 (87.7%) | 2,080 (89.7%) |  |
| **Vital Status** |  |  |  |  | 0.08 |
| Alive | 61 (2.5%) | 54 (2.2%) | 60 (2.5%) | 36 (1.6%) |  |
| Dead | 2,359 (97.5%) | 2,382 (97.8%) | 2,340 (97.5%) | 2,282 (98.4%) |  |

### Receipt of Systemic Therapy

Overall, 46.6% of patients received systemic therapy. There was no significant difference in time to initiation of systemic therapy (0.5-1.4 months) across quartiles. However, patients in the lowest deprivation neighborhoods had a higher rate of systemic therapy receipt (54.0%) compared to those in the highest deprivation neighborhoods (39.4%; p < 0.001). In adjusted analysis with socioeconomic deprivation as a continuous measure, for every unit increase in deprivation the probability of receiving systemic treatment decreased by 1.03% (regression coefficient -0.0103, 95% CI -0.0119 - -0.0086; Figure 1). In adjusted logistic regression, the lowest deprivation quartile had an increased odds of receipt of systemic therapy relative to the highest deprivation quartile (OR 1.93; 95% CI 1.70-2.18, Table 2).

**Figure 1:**
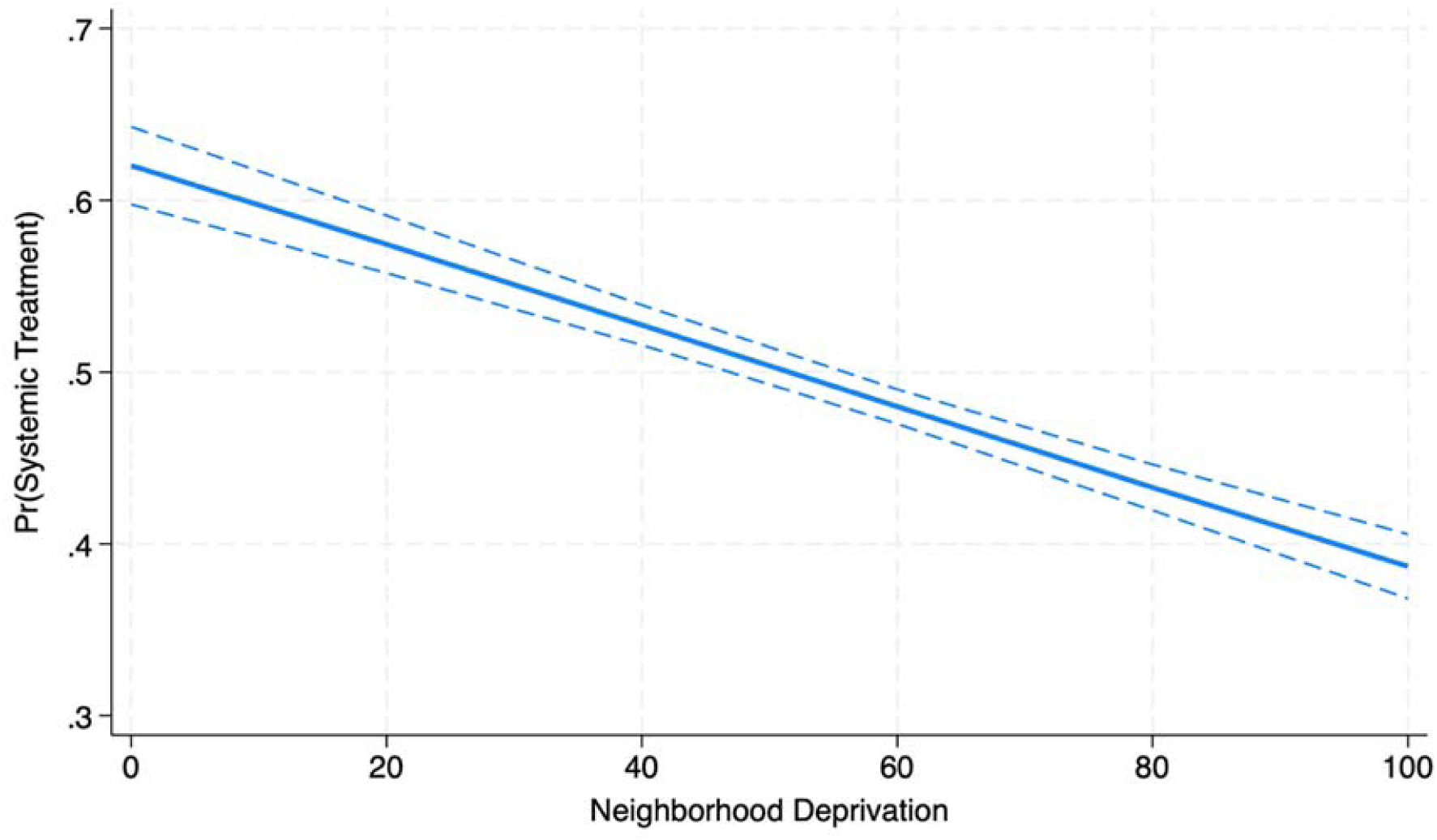
Marginal Plot of the Probability of Systemic Therapy by Neighborhood Socioeconomic Deprivation. Adjusted for age, sex, race, and ethnicity

**Table 2:** Relative Odds and Marginal Probability of Receipt of Systemic Treatment by Neighborhood Deprivation.

| **Neighborhood**<br>**Deprivation (ref:**<br>**Highest (80-100))** | **Odds Ratio (95%**<br>**CI)** | **P value** | **Marginal**<br>**Probability (95%**<br>**CI)** | **P value** |
| --- | --- | --- | --- | --- |
| Q1 Lowest (1-35) | 1.93 (1.70-2.18) | <0.001 | 0.15 (0.12-0.18) | <0.001 |
| Q2 Low (36-57) | 1.52 (1.34-1.71) | <0.001 | 0.10 (0.07-0.12) | <0.001 |
| Q3 High (58-79) | 1.15 (1.02-1.30) | 0.02 | 0.03 (0.01-0.06) | 0.02 |

Adjusted for age, sex, race, and ethnicity

### Overall Survival

The median follow-up time of the cohort was 3.0 months, and the median overall survival was 2.9 months (IQR 1.1-7.6). The 30-day and 90-day mortality of the cohort was 22.2% and 50.1% respectively. By socioeconomic deprivation, there were differences in median overall survival (Table 3 and Figure 2). The median overall survival for the lowest and highest deprivation quartiles were 3.8 months (IQR 3.5-4.1) and 2.4 months (IQR = 2.2-2.5), respectively. When assessed by receipt of systemic therapy, neighborhood socioeconomic deprivation was associated with median overall survival in the cohort receiving systemic treatment (p = 0.01); however, there was no difference in survival in the cohort that did not receive systemic treatment (p = 0.43).

**Figure 2:**
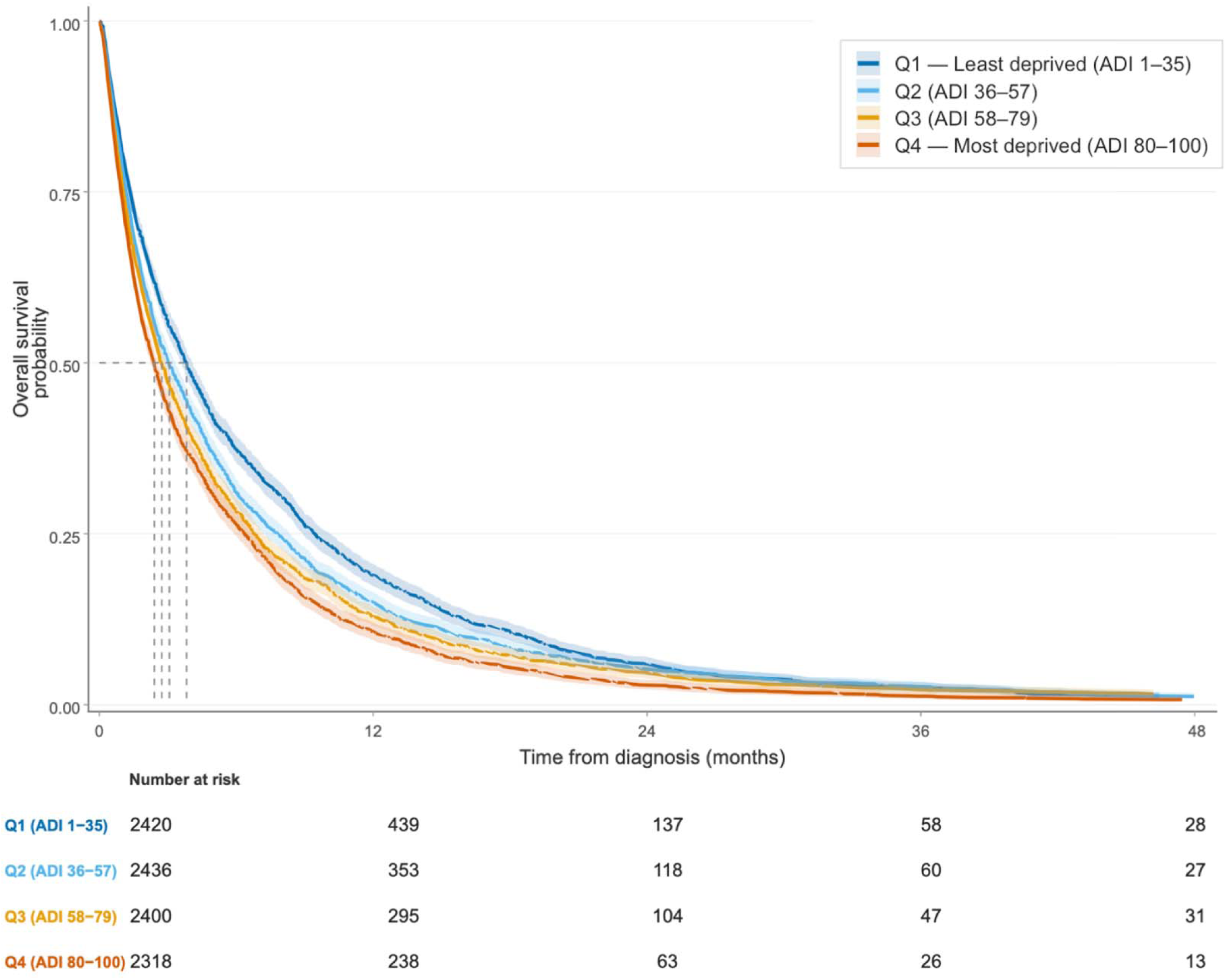
Overall Survival by Socioeconomic Deprivation Quartile IPW-Weighted KM Curve (Log-rank p<0.001)

**Table 3:**
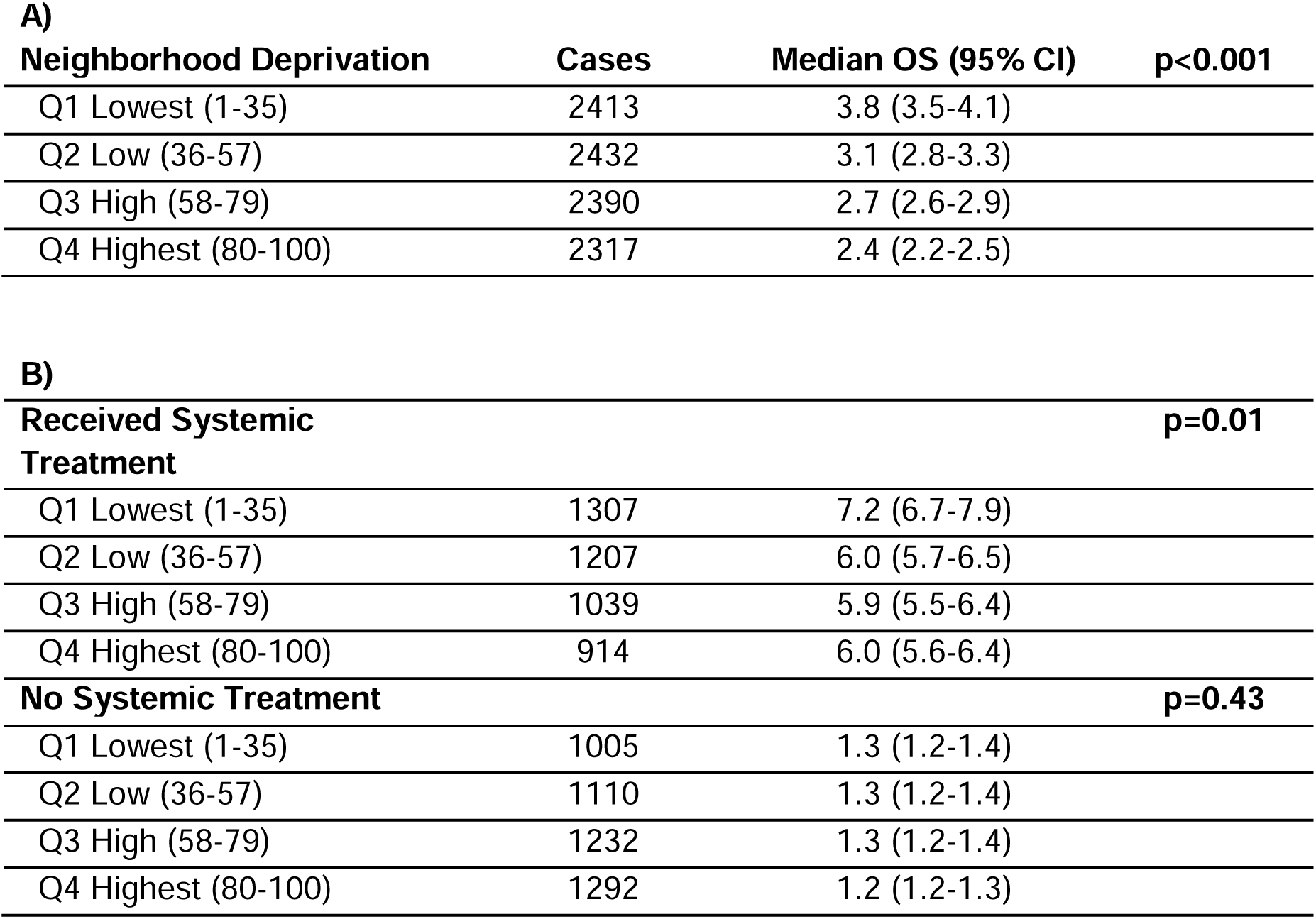
Median Overall Survival by Neighborhood Socioeconomic Deprivation for A) Total Cohort and B) Receipt of Systemic Therapy.

Inverse probability weighting (IPW) achieved adequate covariate balance for 12 of 13 confounders, with absolute standardized mean differences below the pre-specified threshold of 0.10 after weighting (Supplementary Figure 3). The exception was African American race, for which the maximum pairwise adjusted SMD was 0.1005, slightly above the threshold by less than 0.001. Weighted SMDs for all other variables fell well below 0.10 (range 0.001–0.083), representing improvement from pre-weighting SMDs (up to 0.137). Stabilized weights were tightly distributed (Mean 1.00, SD 0.16). In the IPW-weighted pseudo-population, overall survival differed significantly across socioeconomic deprivation quartiles (weighted log-rank p = <0.001; Figure 2).

The estimated marginal hazard ratio for death of the most deprived quartile to the least deprived quartile was 1.32 (95% CI 1.20-1.40), such that under the assumption of sufficient causal adjustment, higher socioeconomic deprivation showed an increased hazard of death (Table 4). Hazard ratios for intermediate quartiles followed a similar gradient, with low deprivation (HR 1.13, 95% CI 1.03-1.20) and high deprivation quartiles (HR 1.21, 95% CI 1.08-1.28).

**Table 4:**
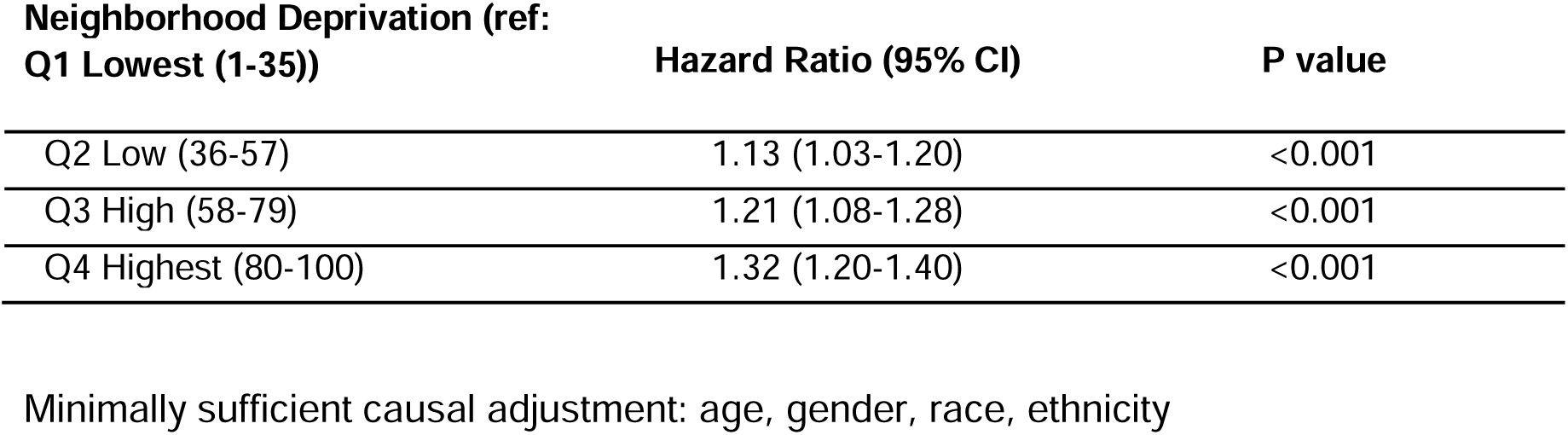
Inverse Probability Weighted Relative Hazard of Death by Neighborhood Socioeconomic Deprivation.

Sensitivity analyses using E-values indicated that an unmeasured confounder would need to be associated with both socioeconomic deprivation and overall survival by a risk ratio of at least 1.69 to fully explain the observed association between the highest and lowest deprivation quartiles, conditional on all measured confounders. Finally, to descriptively assess mediation, sequential covariate adjustment in unweighted Cox models was performed. The hazard ratio for the highest socioeconomic deprivation quartiles compared to the lowest was attenuated from 1.35 (95% CI 1.27-1.43) in the confounder-adjusted model to 1.34 (95% CI 1.27-1.42) after additional adjustment with insurance status and then 1.21 (95% CI 1.13-1.29) after including systemic therapy (Supplemental Table 1). The progressive attenuation of the socioeconomic deprivation and survival association after inclusion of insurance status and receipt of systemic therapy suggest that these variables may partially mediate the relationship between socioeconomic deprivation and overall survival.

## Discussion

In this incidence-based study of metastatic PDAC, most patients did not receive systemic therapy and overall survival for the cohort was poor—the median OS was 2.9 months. Under the stated assumptions, neighborhood socioeconomic deprivation is estimated to have a causal effect on receipt of chemotherapy and overall survival, with higher deprivation leading to less treatment and worse overall survival. Furthermore, there was a dose-response gradient in which each successive quartile of deprivation was associated with an increased hazard of death. Collectively, these findings suggest that socioeconomic disparities impact PDAC treatment and survival.

Few studies have assessed the role of socioeconomic status in metastatic PDAC. This may be due to the challenge of quantifying socioeconomic status, a multidimensional concept that envelops numerous social/eco-social determinants and dynamics such as income, housing, education, etc., all of which may shift over time.^33–35^ With a paucity of individual level socioeconomic data, area-level composite measures of socioeconomic status have emerged as publicly available tools, like the Neighborhood Atlas and Social Vulnerability Index.^36^ These measures help overcome the biases of larger geographic units of measurement, such as zip codes and counties, that are not recommended for socioeconomic status evaluation.^37^ While some studies have assessed the impact of socioeconomic status using the ADI on PDAC treatment and outcomes, these have predominantly been in non-metastatic cohorts.^37–39^

Although the standard of care is systemic treatment with chemotherapy, population-based studies have shown that less than half of metastatic PDAC patients receive systemic treatment, ranging from 31% to 38%. Analysis of the Flatiron database showed a higher first line rate of 74.2%, though it is not incidence-based and is subject to selection bias.^40–42^ For metastatic PDAC patients, prior studies have shown that a higher social vulnerability index score was associated with a lower rate of systemic therapy.^43^ Other studies have suggested insurance status is associated with receipt of treatment.^44^ A single center study suggested that while Yost-based socioeconomic status was associated with race, it was not associated with time to treatment.^45^

For overall survival, population-based studies of metastatic PDAC have shown minimal improvement over several decades despite newer systemic treatment regimens.^4,5,46^ Even among patients in the least deprived quartile, median overall survival was 3.8 months, which is considerably less than the 6.7-11.3 months reported across major phase III trials of first line metastatic PDAC therapies.^47–49^ This disparity likely reflects the highly selected nature of clinical trial populations that tend to be younger, have better performance status, higher SES, are disproportionately White and treated at academic centers; national data showed that only 0.4% of PDAC patients were enrolled in a clinical trial but enrollment was associated with higher median survival for stage IV disease (9.0 months clinical trial vs. 3.8 months non-trial).^50,51^. While clinical trials are crucial for developing novel therapies, the present analysis offers a more generalizable assessment of the real-world burden of metastatic PDAC.

Despite the theoretical advantages, causal inference methods remain uncommon in observational studies using large datasets.^52^ This study sought to assess the effect of socioeconomic deprivation on metastatic PDAC, a question that is not answerable by clinical trial. Nevertheless, there are specific limitations of causal estimation and unmeasured confounding remains a significant barrier. To address confounding from variables not included in the dataset, an E-value analysis was done. This showed that a risk ratio of at least 1.688 and 1.604 (E-value for the lower confidence bound) would be required to explain unmeasured confounding, such as comorbidity. Published estimates of the association between comorbidity burden and overall survival in PDAC populations showed a HR of 1.31-1.55, which fall below this threshold.^53,54^ This suggests that unmeasured confounding by comorbidity alone is unlikely to account for the magnitude of the observed impact of socioeconomic deprivation on overall survival and is consistent with prior studies that have shown that comorbidity only slightly attenuates the SES-survival association.^54^

This study has several additional limitations to consider when interpreting these data. The Florida Cancer Database does not capture certain clinical variables such as patient comorbidities as well as genetic or molecular tumor data, systemic therapy regimen (e.g., type of first line treatment, multi-vs. single agent, and number of cycles), and patient-reported treatment preferences. Additionally, socioeconomic deprivation may not fully reflect individual-level socioeconomic circumstances or changes over time. Further, these data reflect patient outcomes up to 2015 and therefore limit contemporary extrapolation. Finally, the consistency assumption of the causal analysis requires that the effect of a given socioeconomic deprivation quartile is well-defined and does not vary across the specific pathways described in the DAG, which is difficult to fully verify for a complex, multidimensional exposure. The challenge of achieving full balance on African American race across socioeconomic deprivation quartiles perhaps highlights the deeply entangled relationship between race and socioeconomic status. Given that African American race was a strong and partially independent predictor of deprivation quartile assignment may reflect a genuine and irreducible structural relationship, i.e., structural racism, rather than a propensity score misspecification, and can be interpreted as a substantive finding about the nature of race-SES entanglement rather than purely as a methodological limitation.

Despite these drawbacks, this study used rigorous statistical methods to approximate the conditions of a randomized comparison, providing the first attempt to causally estimate these outcomes in an incidence-based metastatic PDAC population. Collectively, these findings provide evidence of a causal effect of socioeconomic deprivation on systemic treatment and overall survival in metastatic PDAC. The magnitude and direction of the survival gradient support the prioritization of socioeconomic equity as a target for PDAC care. Structural interventions, such as patient navigation, transportation assistance, and financial counseling, may represent the most proximal and actionable target for this disparity.

## Data Availability

All data produced are available online at https://fcds.med.miami.edu/inc/welcome.shtml

## Authorship

Each author participated in the work to take public responsibility for appropriate portions of the content as per the guidelines of the International Committee of Medical Journal Editors (ICMJE). All authors made substantial contributions to the conception or design of the work; or the acquisition, analysis, or interpretation of data for the work; AND drafting the work or revising it critically for important intellectual content; AND final approval of the version to be published; AND agreement to be accountable for all aspects of the work in ensuring that questions related to the accuracy or integrity of any part of the work are appropriately investigated and resolved.

