## Supplementary figures and images for "The impact of neighborhood socioeconomic deprivation on metastatic pancreatic cancer treatment and survival: An incidence-based, causally-structured observational study"

### Supplemental Figure 1

**Supplemental Figure 1: Cohort Selection**

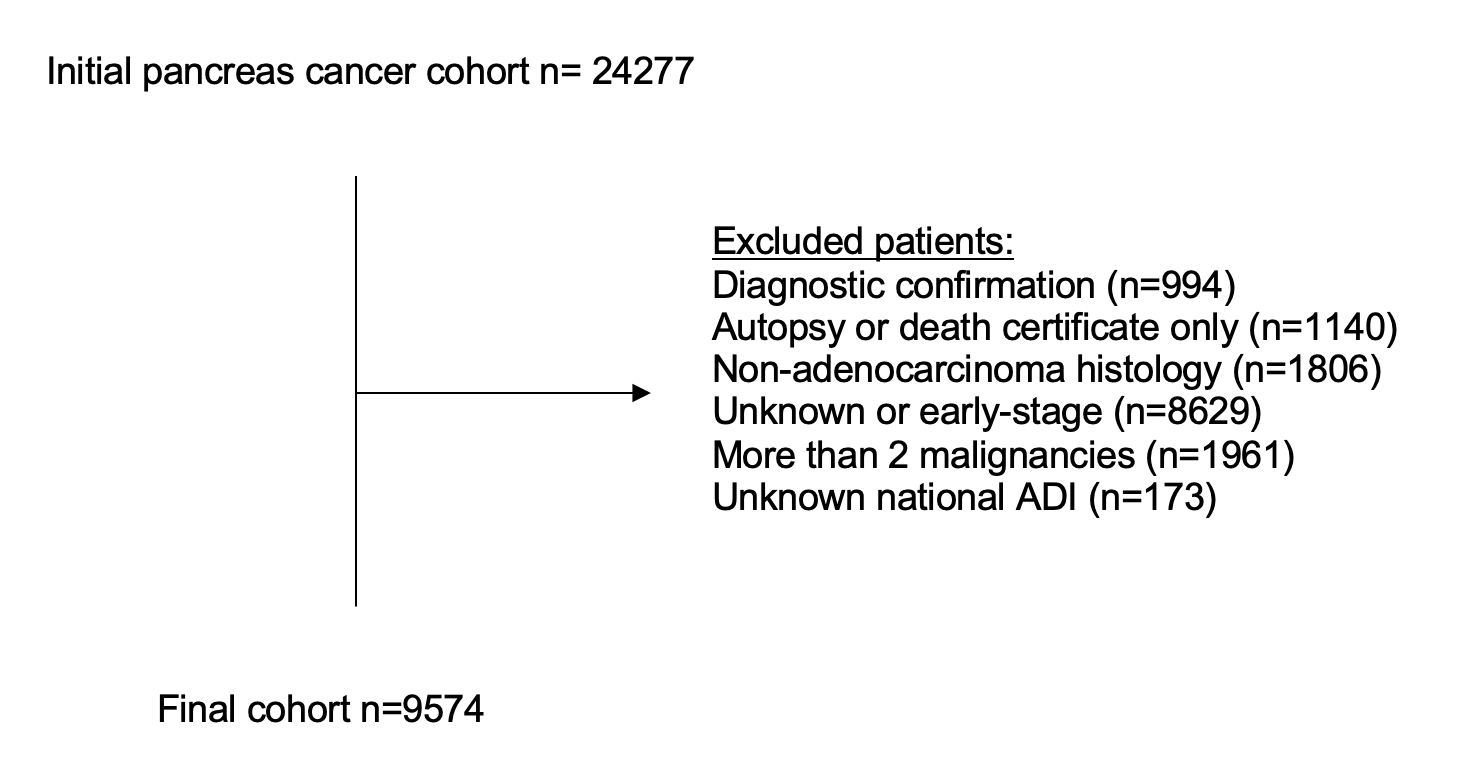

### Supplemental Figure 3

**Supplemental Figure 3: IPW Covariate Balance**

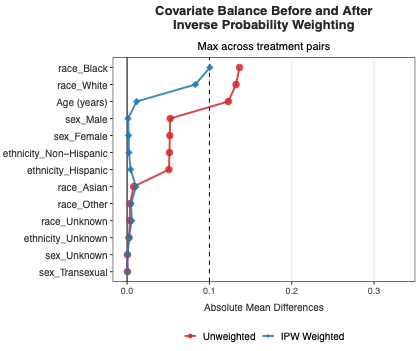
