## Supplemental Figure 2 for "The impact of neighborhood socioeconomic deprivation on metastatic pancreatic cancer treatment and survival: An incidence-based, causally-structured observational study"

**Supplemental Figure 2: Directed acyclic graph of exposure, socioeconomic deprivation, and outcome, overall survival for total effect**

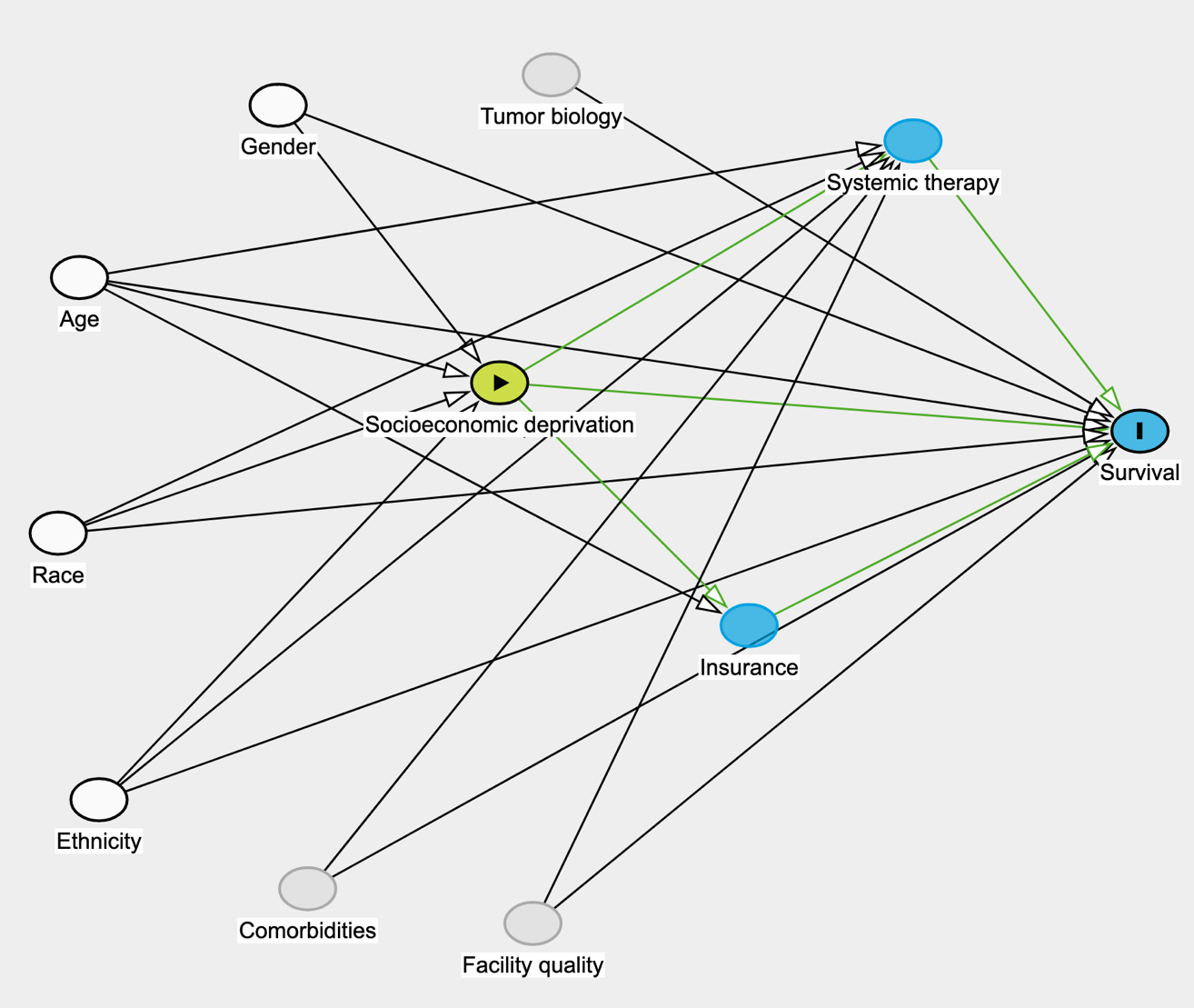
 Factors required for minimal sufficient adjustment: age, ethnicity, race, and sex.

**Legend:**

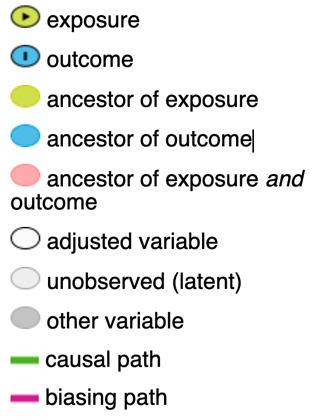
