## Supplemental Table 1 for "The impact of neighborhood socioeconomic deprivation on metastatic pancreatic cancer treatment and survival: An incidence-based, causally-structured observational study"

**Supplemental Table 1: Relative Hazard of Death by Neighborhood Socioeconomic Deprivation**

| **Neighborhood Deprivation (ref: Q1 Lowest (1-35))** | **Model 1**  **Hazard Ratio (95% CI)** | **P value** | **Model 2**  **Hazard Ratio (95% CI)** | **P value** | **Model 3**  **Hazard Ratio (95% CI)** | **P**  **value** |
| --- | --- | --- | --- | --- | --- | --- |
| Q2 Low (36-57) | 1.13 (1.03-1.20) | <0.001 | 1.13 (1.07-1.20) | <0.001 | 1.11 (1.04-1.17) | 0.001 |
| Q3 High (58-79) | 1.22 (1.15-1.29) | <0.001 | 1.22 (1.15-1.29) | <0.001 | 1.12 (1.06-1.19) | <0.001 |
| Q4 Highest (80-100) | 1.35 (1.27-1.43) | <0.001 | 1.34 (1.27-1.42) | <0.001 | 1.21 (1.13-1.29 | <0.001 |

Model 1 adjustment: age, gender, race, ethnicity

Model 2 adjustment: age, gender, race, ethnicity, insurance status

Model 3 adjustment: age, gender, race, ethnicity, insurance status, and receipt of systemic treatment
